# A study on the relationship between BCG vaccination and Covid-19 prevalence: Do other confounders warrant investigation?

**DOI:** 10.1101/2020.05.06.20093138

**Authors:** Richard M. Mariita, Jonathan M. Musila

## Abstract

The Covid-19 pandemic, which originated from Wuhan, Hubei province, China, and quickly spread to the rest of the globe is caused by SARS-CoV-2, a single-stranded RNA virus. Preliminary data suggest a relationship between the BCG vaccine and the prevalence of Covid-19. The BCG vaccine is used in the prevention of tuberculosis, a disease that is most prevalent in developing countries. To determine the potential protective role of BCG vaccination, this study investigated the occurrence of Covid-19 and the relationship between the spread of Covid-19 in countries that offer BCG vaccination and those that do not. The study also performed a phylogenetic analysis of the strains involved in the Covid-19 outbreak from the representative countries. To achieve the objectives, the study utilized publicly available data on population size, vaccination coverage, and Covid-19 cases. Phylogenetic analysis was used to determine if some SARS-CoV-2 strains were more prevalent than others. The study revealed a significant negative trend between countries that offer the BCG vaccine to the general population and the reported cases of Covid-19. The study proposes future molecular and immunological analyses to determine the potential role of BCG vaccination in protection against Covid-19. This will determine if BCG has antiviral properties, with the possibility of recommending it for widespread use if supported by scientific data.

## Introduction

Over the last century, mankind has faced numerous pandemics associated with the emergence of new microorganisms, the latest being the Covid-19 pandemic caused by severe acute respiratory syndrome coronavirus 2 (SARS-CoV-2) (Andersen et al., 2020). As of 30th April 2020, this virus was present in 212 countries and territories (Kang et al., 2020). Pandemics occur when novel microorganisms, usually viruses emerge as a result of mutations capable of coding for new potentiator biomolecules such as the spike glycoprotein by SARS-CoV-2 (Giovanetti et al., 2020). Consequently, the new machinery may enable the pathogen to jump between hosts, for instance from bats to pangolins and then from the intermediate host to humans (Andersen et al., 2020). Prompt data collection of the novel organisms is critical to understanding behavioral and infectious trends of these pathogens, and in disease mitigation by generating relevant antimicrobial agents (AMA) and vaccines. Efforts to generate effective AMA for novel organisms are challenged by limited understanding of the infectious agent, and hence monitoring basic trends becomes crucial. Following the spread of Covid-19, there have been mixed and unprecedented trends, and varying findings are still accumulating. Interestingly, for instance, countries that immensely practice BCG vaccination against tuberculosis (TB), caused by a bacterium (*Mycobacterium tuberculosis*) that has a deleterious respiratory effect (Curtis et al., n.d.), have generally reported low infection rates which has led to suggestions that BCG vaccination could be slowing or helping in the protection against Covid-19 (Curtis et al., n.d.). Previous studies have demonstrated that the BCG vaccine has non-specific beneficial effects (Curtis et al., n.d.) (Uthayakumar et al., 2018), including increased protection from a variety of respiratory infectious diseases (Prentice et al., 2015). Some of these studies have revealed possible epigenetic changes that subsequently regulate cytokine production (Uthayakumar et al., 2018) (Arts et al., 2018). Coincidentally, most Covid-19 victims die due to unregulated cytokine production.

In this study, we sought to find a correlation between BCG vaccination coverage and Covid-19 virus cases to determine if indeed that pattern arises. It has also been argued that because of variation in geographical positioning and comorbidities of the affected people (Guan et al., 2020a), pandemics tend to cluster and generally impact different parts of the world to varying extents (https://coronavirus.jhu.edu/). As such, the novel Covid-19 virus is no exception with early data showing variation in distribution to different regions of the world (Ensheng et al., 2020). Herein, we discuss how the impact and the behavioral trend of the Novel Covid-19 virus might have been influenced by geographical positioning, and immunization of the populace especially against TB. The current study aimed to evaluate the relationship between BCG vaccination and coronavirus isolated strains, thus the prevalence of Covid-19.

### Methodology

Data on Covid-19 pandemic cases was obtained from The Johns Hopkins Coronavirus Resource Center (https://coronavirus.jhu.edu/) and Google News (https://news.google.com/covid19). BCG vaccination data for the representative countries was obtained from BCG Atlas (http://www.bcgatlas.org/). Data analysis and graphing were performed using GraphPad Prism version 8 (https://www.graphpad.com/). Population data for confirmation of infected ratio was obtained from https://blog.lazd.net/coronaglobe/. Publicly available genomic data was mined from Global Initiative on Sharing All Influenza Data (GISAID) (https://www.gisaid.org/) (Elbe and Buckland-Merrett, 2017) and https://www.ncbi.nlm.nih.gov/. Phylogenetic analysis was done using the Tamura-Nei model on MEGA X (Kumar et al., 2018) and Nextstrain (https://nextstrain.org/) was used to generate phylogenetic trees for clades.

### Results and Discussion

The Covid-19 pandemic has had a global impact (Fig. 1) and a high fatality rate. By the end of April 2020, there were 3,303,296 confirmed cases and 235,290 deaths globally. Further emerging in silico predictions reflect even more infections and fatalities. However, European countries and the USA have reported the highest number of cases per one million people (Ensheng et al., 2020). India and the source of the pandemic, China, had lower cases per one million people compared to the USA, Italy, Spain, Netherlands, and UK, even though they are the most populated countries in the world (Table 1). African countries under study had hardly reported high cases of Covid-19 by the date this article was written (end of April 2020). However, there was an initial fear that due to weaker healthcare systems, Chinese expatriates (about 2 million Chinese nationals live and work in Africa), and the associated increased travel between China and Africa for education, business and leisure, the infection rates could be higher (Kapata et al., 2020).

**Fig. 1:**
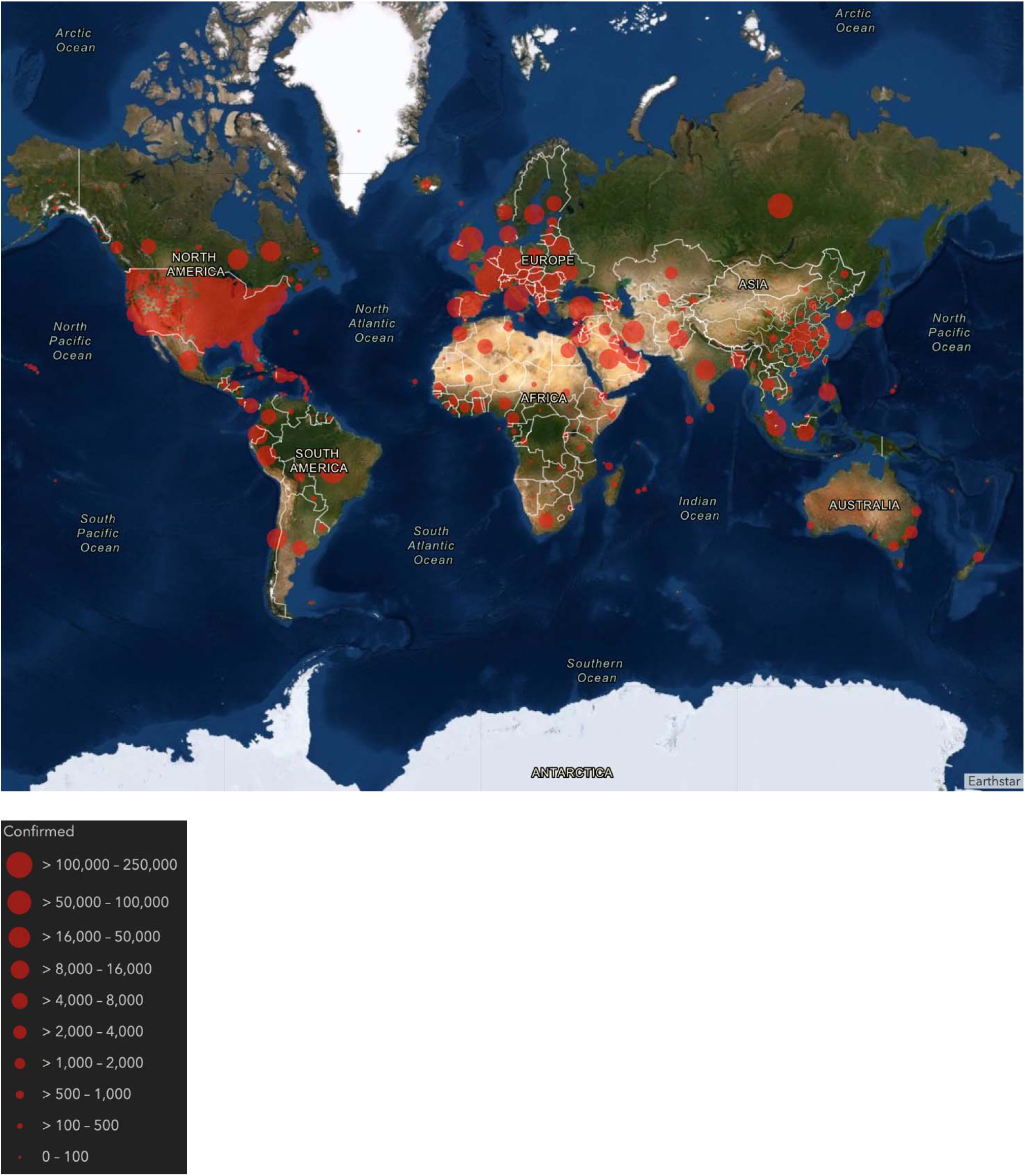
Globally confirmed cases of Covid-19 pandemic disease

**Table 1:**
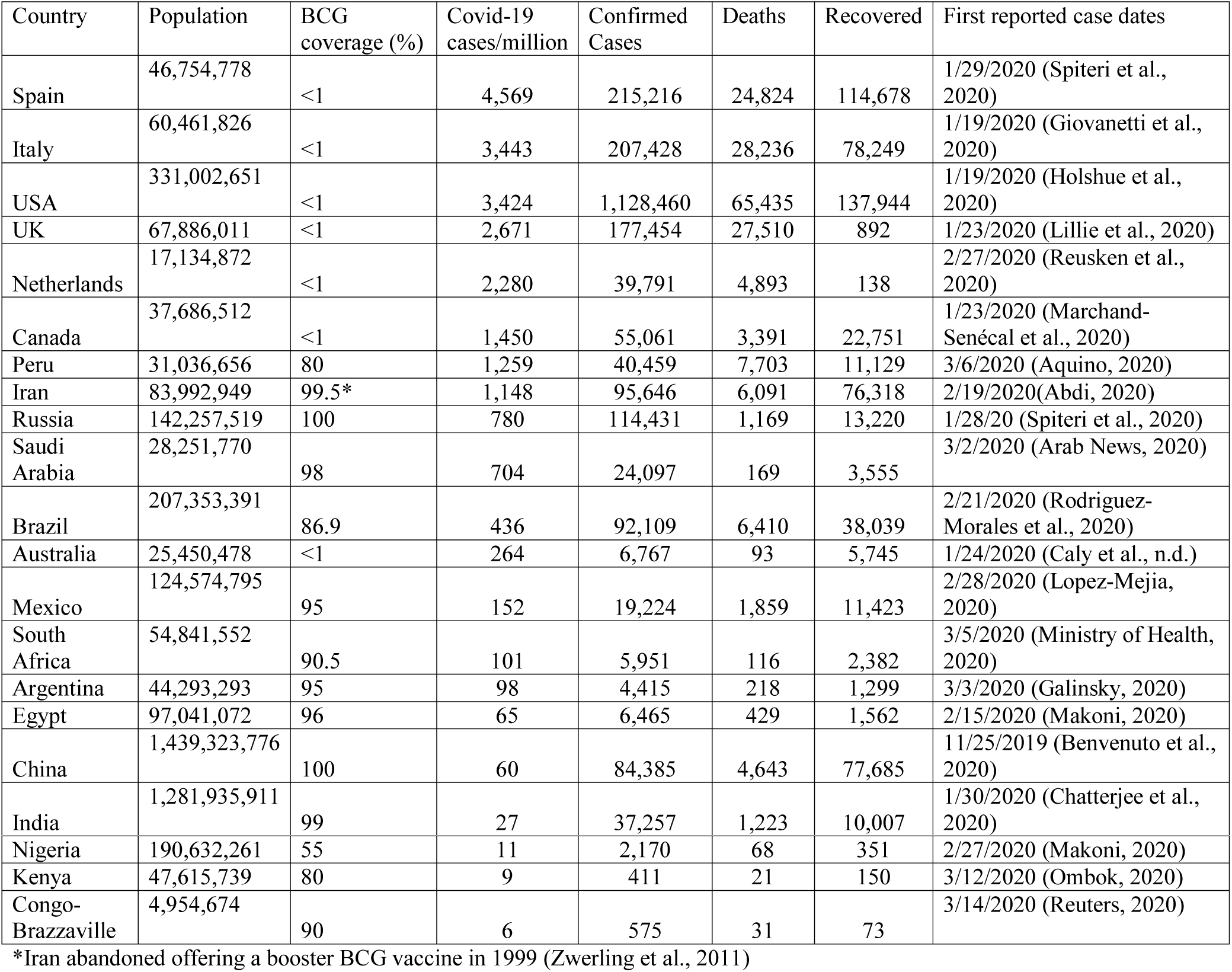
Proportion of Covid-19 cases, BCG vaccination coverage and affected population as of end of April 2020

Coincidentally, countries with lower cases per million people had one thing in common: BCG vaccination (Fig. 2). Interestingly, Iran, a country that abandoned offering a booster BCG vaccine in 1999 (Zwerling et al., 2011), registered high cases of Covid-19 (Fig 3). It is, however, hard to postulate the impact that may have had on Covid-19 incidences. The low number of cases in China could be due to the underlying effects of BCG vaccination and/or massive efforts by the government to contain the virus. Overall, countries with low TB incidences (mostly western countries) that do not administer the BCG vaccine to the general population had the highest Covid-19 cases per million people. Countries such as England stopped the BCG vaccination in 2005 (Fine, 2005) whereas Australia abandoned its program in the 1980s due to a reduction of tuberculosis incidences (Zwerling et al., 2011). India has an expansive BCG vaccination program (Zwerling et al., 2011). The country registered 27 cases per million people by the end of April 2020. Additionally, India’s reported first case was on 1/30/2020 (Table 1), and by April 5, 2020, the 1.3 billion people in India were put into lockdown. This precaution could have added to the containment of the virus spread. An even more interesting contrast was the low number of Covid-19 cases in the United States’ southern neighbor, Mexico; while Mexico reported (152 cases per million by the end of April 2020, the USA had 3,424 cases per million by the same date. A similar trend was seen between the USA and Argentina, a South American state with high BCG vaccination coverage.

**Fig. 2:**
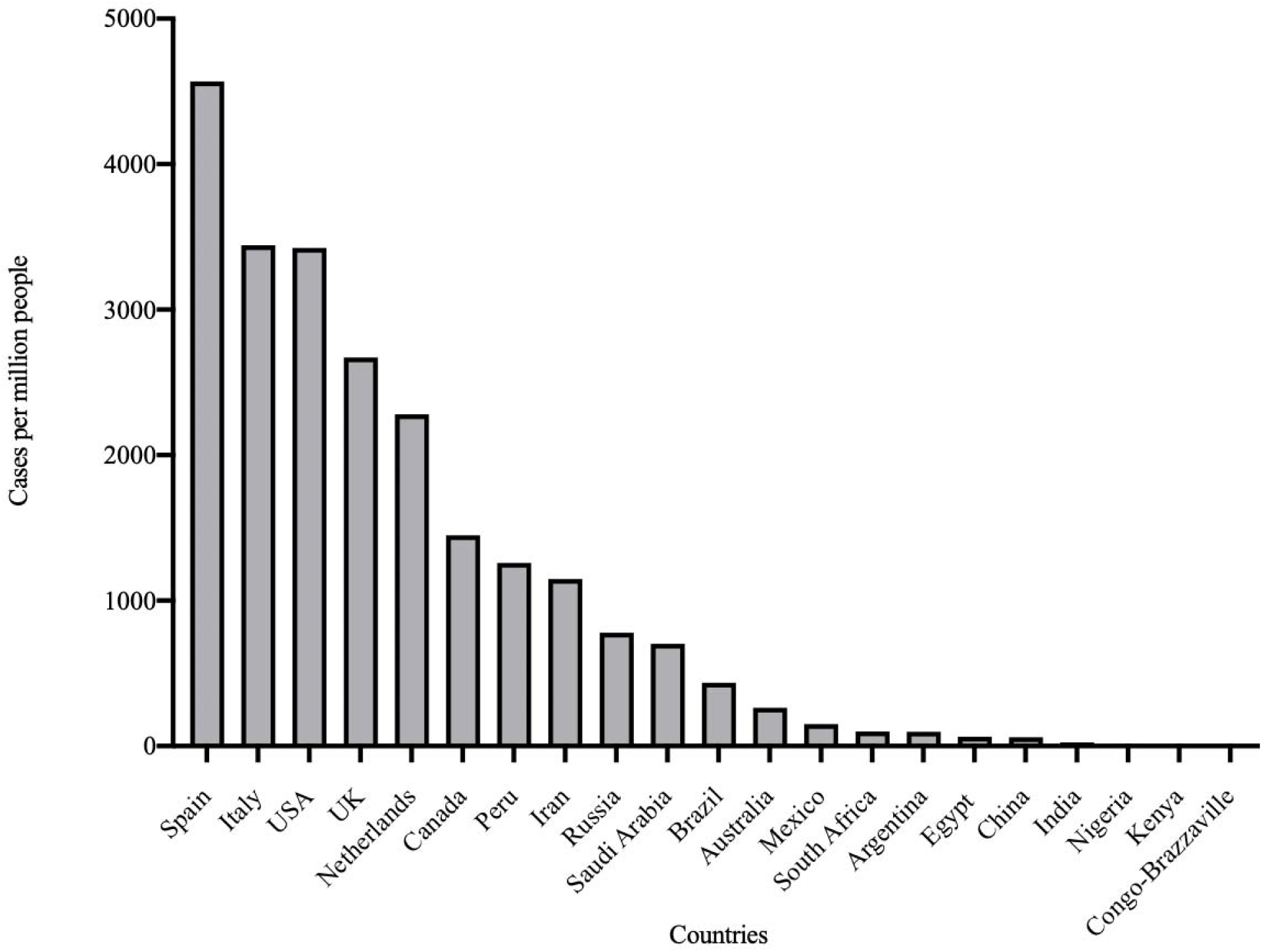
Cases per million for representative countries revealed high cases in developed countries

**Fig. 3:**
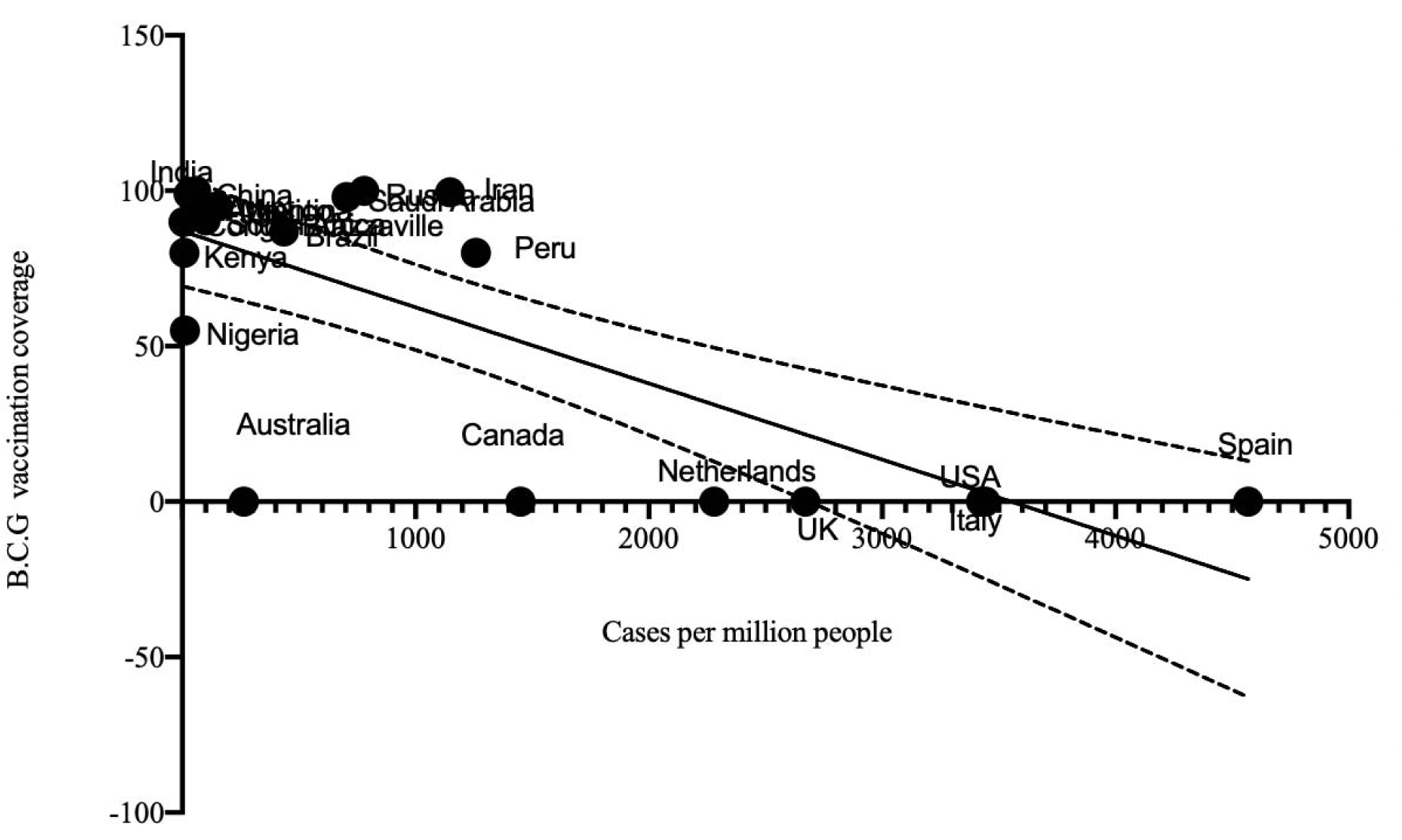
Representative countries with BCG vaccination programs had lower reported cases of Covid-19 (with 95% asymptotic confidence bands)

The BCG vaccination coverage had a negative correlation with reported Covid-19 cases (*p*<0.0001, R^2^=0.5707). Regression analysis revealed a strong association between low numbers of Covid-19 cases and BCG vaccination (mean value of 75.54%) (Fig. 3). However, high numbers of Covid-19 cases cannot be explained by the lack of BCG vaccination alone. There are other risk factors such as comorbidity, age, and socio-economic factors including living conditions (Guan et al., 2020b) (Yang et al., 2020). Phylogenetic analysis using genomic data reveals that some countries that were highly impacted shared strains with the same ancestral nodes with those with less Covid-19 reported cases (Fig. 4). This could mean shared strains alone may not determine cases, but BCG, which is shared among those reporting lower cases could be one factor, among other confounders. Further analysis revealed clade A2a was most common in Africa, North and South America, Europe, and Asia (Fig. 5). Clade A2a was curiously the most common in New York City’s most affected areas of Manhattan, Brooklyn, Bronx, and Westchester (Gonzalez-Reiche et al., 2020). The populations in New York City boroughs and its neighborhoods are considerably high. But these are not as poorly sanitized regions as slums in developing countries (Corburn and Hildebrand, 2015). Further, overcrowding (such as Kenya’s Kibera slums which has 30,000 persons per square mile) (Njuguna et al., 2013), poor infrastructure, and inadequate social amenities (Kamau and Njiru, 2018), would have led to a catastrophic situation. Further, looking at Saudi Arabia and Egypt (both with >95% BCG coverage) emphasizes the possible existence of other confounders besides BCG vaccination (Table1).

**Fig. 4:**
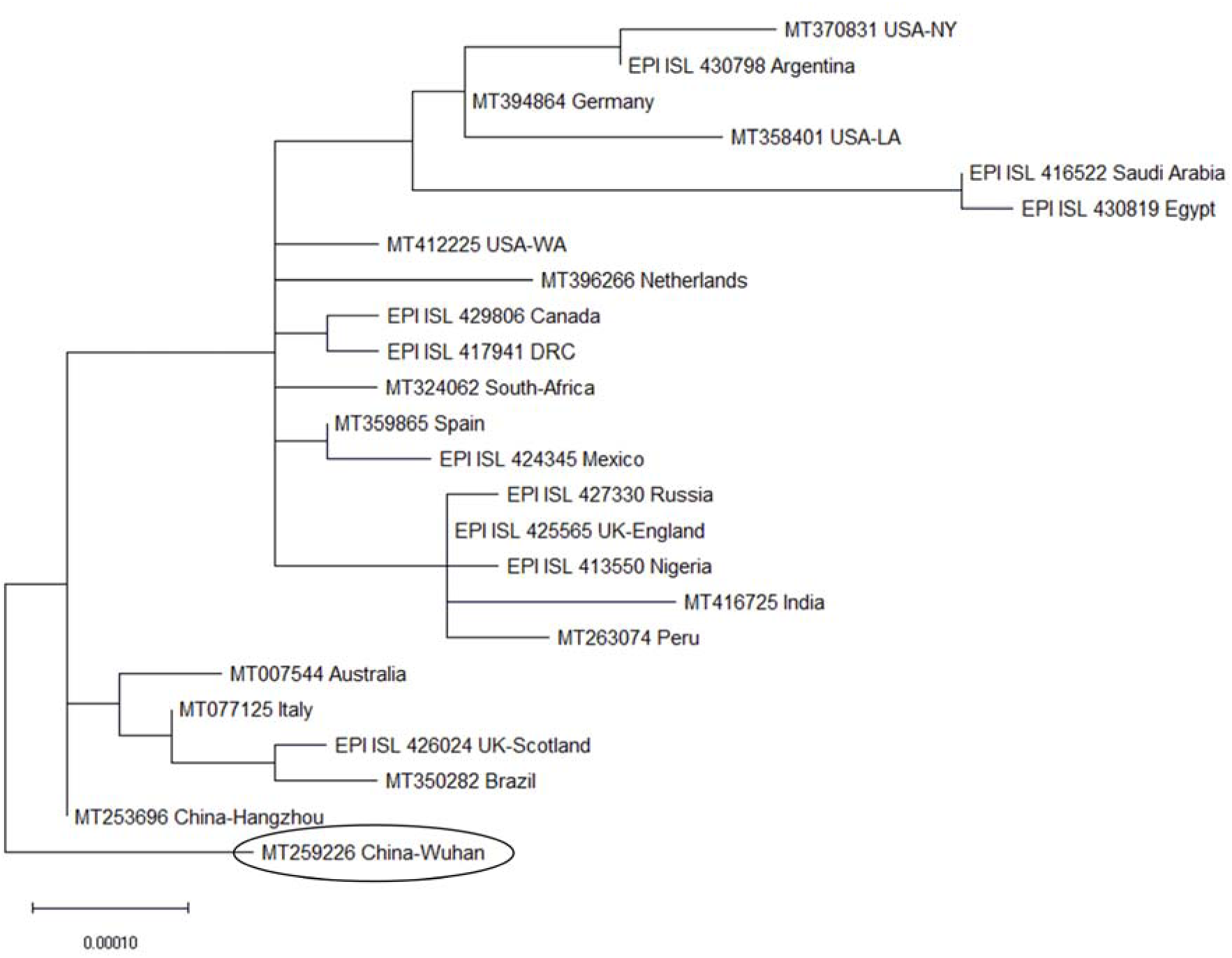
Phylogenetic network of 24 SARS-CoV-2 genomes from most of the representative countries. The evolutionary history was inferred by using the Maximum Likelihood method and the Tamura-Nei model. The tree with the highest log likelihood (−41348.44) is shown and was rooted in SARS-COV-2 nucleotide sequence from China-Wuhan (encircled). Initial tree(s) for the heuristic search were obtained automatically by applying Neighbor-Join and BioNJ algorithms to a matrix of pairwise distances estimated using the Tamura-Nei model, and then selecting the topology with superior log likelihood value. The tree is drawn to scale, with branch lengths measured in the number of substitutions per site. This analysis involved 24 nucleotide sequences. There was a total of 29911 positions in the final dataset. Evolutionary analyses were conducted in MEGA X (Kumar et al., 2018).

**Fig. 5:**
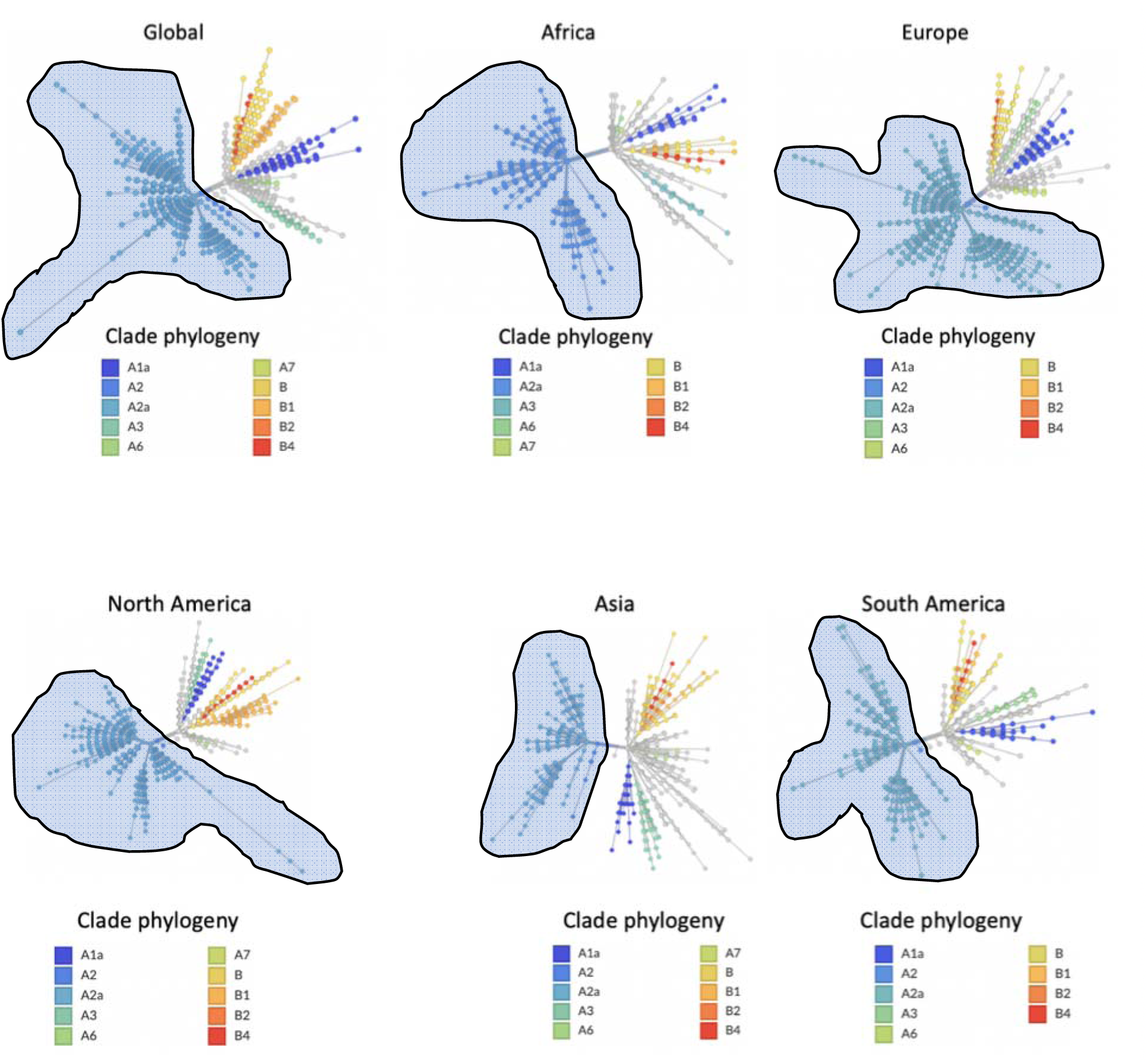
Phylogeny showed that clade A2a (indicated in freeform) was the most common in representative geographical locations

Countries which have an immense BCG vaccination practice continue to manifest low levels of Covid-19 spread. Although BCG induced changes have been found to correlate with protection against experimental viral infection (Arts et al., 2018), it is not precisely clear whether there is such protection against coronaviruses. Consequently, we assume that other factors such as but not limited to weather, sanitation, comorbidity, continuous evolution of the virus, and the intensity of Covid-19 testing play a role, confounders which warrant further individual and/or collective investigation.

## Conclusion

We assessed the relationship between BCG vaccination and the number of Covid-19 cases. The study revealed a significant negative trend between countries that offer BCG vaccinations to the general population and reported cases of Covid-19. Whereas the study established a correlation, other factors could have contributed to observed differences including but not limited to, test capabilities, demographics, and disease burden. To elucidate direct antiviral benefits of BCG vaccination, molecular analysis to determine the role of BCG vaccination and potential protection against coronaviruses should be investigated. If BCG is found to have coronavirus protection benefits, the clinical need for a BCG vaccination should be evaluated in countries that do not administer BCG vaccination, particularly vulnerable and easily exposed population groups, such as, the elderly, those with comorbidities and healthcare workers. Moreover, evidence of protection against viral respiratory infectious agents should underscore the need for developing countries to continue the administration of BCG vaccination promptly to offer continued beneficial effects on its population.

## Data Availability

The nucleotide sequences utilized in phylogenetic analysis (Fig. 5) were obtained from https://www.ncbi.nlm.nih.gov/ (Accession number starts with MT) and https://www.gisaid.org/ (Accession number starts with EPI_ISL_) and includes the following: MT412225 (USA-WA), MT370831 (USA-NY), MT358401 (USA-LA), EPI_ISL_429806 (Canada), EPI_ISL_424345 (Mexico), EPI_ISL_430798 (Argentina), MT350282 (Brazil), MT263074 (Peru), EPI_ISL_425565 (England), EPI_ISL_426024 (UK-Scotland), MT394864 (Germany), MT396266 (Netherlands), MT359865 (Spain), MT077125 (Italy), EPI_ISL_417941 (DRC), EPI_ISL_430819 (Egypt), EPI_ISL_413550 (Nigeria), MT324062 (South-Africa), EPI_ISL_427330 (Russia), EPI_ISL_416522 (Saudi-Arabia), MT416725 (India), MT259226 (China-Wuhan), MT253696 (China-Hangzhou) and MT007544 (Australia). The alignment sequences are in the supporting information and can be accessed via https://doi.org/10.6084/m9.figshare.12246302.v1. Evolutionary analyses were conducted in MEGA X (Kumar et al., 2018).

## Data Availability

The alignment sequences are in the supporting information and can be accessed via provided links.

https://doi.org/10.6084/m9.figshare.12246302.v1.

## Acknowledgment

We gratefully acknowledge the authors, originating and submitting laboratories of the sequences from GISAID’s EpiFlu™ and Nextstrain Databases. Authors also recognize Coronavirus Resource Center at Johns Hopkins University, Google’s COVIS-19 news, and BCG atlas for the provision of data to the public. Authors lastly thank Philip Alabi (Brown University, Rhode Island), Rosemary Nyamboya (St George’s University Hospital (London, UK), Courtney Mariita (Troy Prep Charter Schools, New York) and Michael Ochien’g (Pontificia Universidad Comillas, Madrid, Spain) and Jessica Gilpin (Vanderbilt University, Tennessee) for their insights and feedback.

## Authors contribution

Both RMM and JMM conceived, designed the study, drafted the manuscript, and contributed to the writing and finalizing of the manuscript.

